# Diagnostic Performance of Self-Supervised Foundation Models for Intraoperative Quantification of Hepatic Macrovesicular Steatosis

**DOI:** 10.1101/2025.09.16.25335833

**Authors:** Shunsuke Koga, Anjani Guda, Yujie Wang, Aarush Sahni, Jiahui Wu, Alyssa Rosen, Jaxson Nield, Nilan Nandish, Krunal Patel, Haviva Goldman, Chamith S. Rajapakse, Selemon Walle, Kristen Stashek, Rashmi Tondon, Zahra Alipour

## Abstract

**Introduction:** Accurate intraoperative assessment of macrovesicular steatosis in donor liver biopsies is critical for transplantation decisions but is often limited by inter-observer variability and freezing artifacts that can obscure histological details. Artificial intelligence (AI) offers a potential solution for standardized and reproducible evaluation. To evaluate the diagnostic performance of two self-supervised learning (SSL)-based foundation models, Prov-GigaPath and UNI, for classifying macrovesicular steatosis in frozen liver biopsy sections, compared with assessments by surgical pathologists.

**Methods:** We retrospectively analyzed 131 frozen liver biopsy specimens from 68 donors collected between November 2022 and September 2024. Slides were digitized into whole-slide images, tiled into patches, and used to extract embeddings with Prov-GigaPath and UNI; slide-level classifiers were then trained and tested. Intraoperative diagnoses by on-call surgical pathologists were compared with ground truth determined from independent reviews of permanent sections by two liver pathologists. Accuracy was evaluated for both five-category classification and a clinically significant binary threshold (<30% vs. ≥30%).

**Results:** For binary classification, Prov-GigaPath achieved 96.4% accuracy, UNI 85.7%, and surgical pathologists 84.0% (*P* = .22). In five-category classification, accuracies were lower: Prov-GigaPath 57.1%, UNI 50.0%, and pathologists 58.7% (*P* = .70). Misclassification primarily occurred in intermediate categories (5%–<30% steatosis).

**Conclusions:** SSL-based foundation models performed comparably to surgical pathologists in classifying macrovesicular steatosis, at the clinically relevant <30% vs. ≥30% threshold. These findings support the potential role of AI in standardizing intraoperative evaluation of donor liver biopsies; however, the small sample size limits generalizability and requires validation in larger, balanced cohorts.

## Introduction

Orthotopic liver transplantation is the standard treatment for end-stage liver disease, but outcomes depend significantly on donor organ quality. Among various histopathologic factors, macrovesicular steatosis strongly influences graft viability, with increasing levels of steatosis correlated with increased risks of primary non-function and early allograft dysfunction.^1–3^ While donor livers with mild macrovesicular steatosis (<30%) generally show comparable outcomes to non-steatotic grafts,^4^ moderate-to-severe steatosis (≥30%) markedly compromises graft survival and overall transplant success.^5^ Recent large-scale analyses revealed that livers with macrovesicular steatosis ≥31% have significantly reduced transplant utilization rates, with approximately 55% of these organs discarded due to elevated concerns regarding graft failure.^3^ Despite these risks, steatotic donor livers remain a valuable resource, particularly as their prevalence rises alongside increasing obesity and metabolic syndrome rates. Therefore, precise and standardized assessment of macrovesicular steatosis is critical to optimizing organ utilization while mitigating transplant risks.

In current practice, intraoperative frozen section evaluation of donor liver biopsies is widely used to assess steatosis and guide real-time transplant decisions.^6–8^ However, this approach has inherent limitations, including freezing artifacts that obscure tissue architecture and significant inter-observer variability in steatosis estimation. Additionally, there is considerable variability among pathologists, including fundamental disagreements regarding concepts such as microvesicular steatosis.^9^ This variability highlights the limitations of relying solely on pathologist-based assessments and underscores the need for more objective and standardized methods. Furthermore, intraoperative frozen sections are often evaluated by on-call pathologists who infrequently assess liver biopsies and thus face challenges interpreting freezing artifacts. These factors contribute to discrepancies between frozen and permanent section assessments, leading to potential misclassification of graft suitability.^10^ Overestimation of steatosis may result in unnecessary organ discard, while underestimation increases the risk of transplanting marginal grafts with compromised function. Objective and standardized assessment methods, such as artificial intelligence-based analysis trained to recognize steatosis and freezing artifacts, may decrease subjective variability, reducing unnecessary organ discard and the risk associated with marginal graft transplantation.

AI-based digital pathology offers promising solutions to these challenges.^11–17^ Supervised approaches such as convolutional neural networks (CNNs) require large annotated datasets, while self-supervised learning (SSL) methods can learn effectively from unlabeled images.^13,18^ Both approaches can rapidly and consistently analyze digitized histology images, reduce subjective variability, and provide quantitative measures. Previous CNN-based studies for steatosis quantification in donor liver biopsies have shown strong correlations (r = 0.85, ICC = 0.85) with expert annotations, highlighting their potential to outperform intraoperative pathologists assessments.^19^ Another CNN-based study reported independent associations between AI-based measurements and early allograft dysfunction, with improved predictive calibration compared to manual evaluation.^20^ Integrating AI models into intraoperative frozen-section workflows could therefore enhance the accuracy and reliability of donor liver assessments, facilitating more informed decisions regarding organ acceptance.^21^

In this study, we evaluated two recently developed SSL-based foundation models, Prov-GigaPath ^22^ and UNI,^23^ for assessing macrovesicular steatosis in 131 frozen liver biopsy specimens. We compared their performance with intraoperative frozen-section diagnoses from pathologists, using consensus ground truth determined from permanent-section, evaluated by two experienced liver pathologists.

## Materials and Methods

### Cohort

This retrospective study included 131 donor liver biopsy specimens from 68 donors, obtained during organ procurement procedures performed at the Hospital of the University of Pennsylvania (Philadelphia, Pennsylvania) between November 2022 and September 2024. All biopsy specimens were collected in collaboration with the Gift of Life Donor Program.^24^ The hospital follows a subspecialty sign-out model, and donor liver biopsies are interpreted on frozen section by the on-call service. During this period, a total of nineteen board-certified anatomic pathology attendings served on the on-call frozen-section service and evaluated donor liver biopsies contemporaneously during organ procurement. The overall workflow of the study is summarized in **Figure 1**. The study protocol was approved by the Institutional Review Board at the Hospital of the University of Pennsylvania.

**Figure 1.**
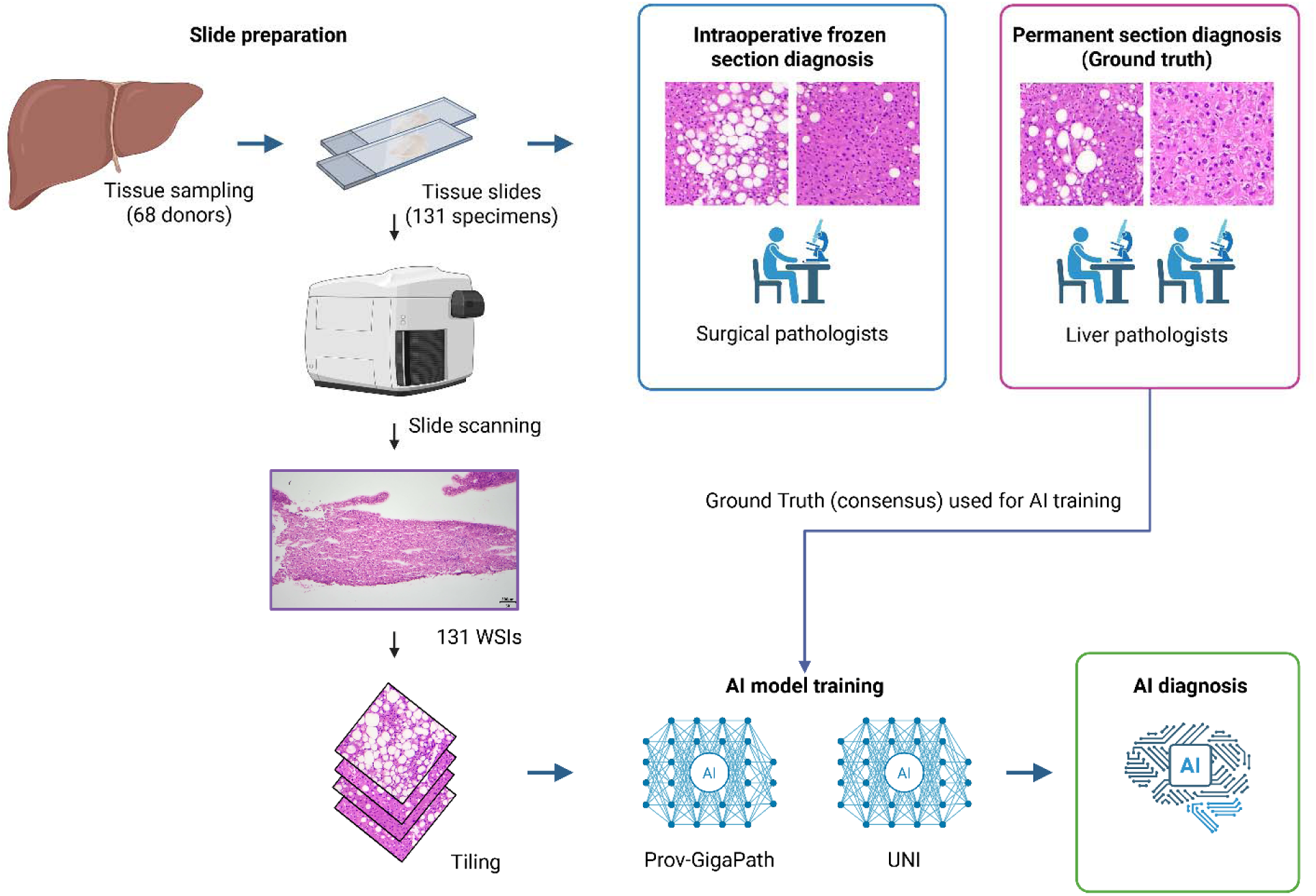
Workflow of AI-based evaluation and pathologist assessment of donor liver biopsies. Liver biopsies are obtained from 68 donors, yielding 131 specimens. Slides are hematoxylin & eosin (H&E)-stained and scanned to generate 131 whole-slide images (WSIs), which are tiled into image patches used for training and testing artificial intelligence (AI) models. Intraoperative frozen section diagnoses (H&E-stained) are made by on service or on call surgical pathologists as part of routine clinical workflow. Permanent H&E-stained sections are independently reviewed by two liver pathologists specifically for this study to establish the consensus ground truth. AI-based diagnoses are generated using two foundation models, Prov-GigaPath and UNI.

### Pathological Assessment and Ground Truth Determination

Macrovesicular steatosis was defined as the presence of large lipid droplets within hepatocytes that displace the nucleus, consistent with definitions widely adopted by liver transplant pathologists.^25^ Microvesicular steatosis was not assessed in this study due to its limited established clinical relevance in transplantation decisions, inconsistent usage of the term among pathologists, and lack of standardized definitions and assessment criteria across institutions.^9^ Steatosis categories were determined according to the Gift of Life Donor Program’s standard pathology evaluation form, as described previously, and classified into five categories: <5%, 5% to <10%, 10% to <20%, 20% to <30%, and ≥30%. These categories were applied for both intraoperative frozen-section assessments and permanent-section reviews.

In December 2024, two liver pathologists (RT and ZA), both with fellowship training in medical liver, independently established the ground truth by reviewing permanent hematoxylin & eosin (H&E) sections, without access to the frozen-section diagnoses at the time of review.

When discrepancies occurred between the two pathologists, the slides were jointly reviewed with a third GI pathologist (KS) to reach consensus.

### Digital Pathology and Feature Extraction

Whole-slide images (WSIs) of frozen liver biopsies were digitized using a high-throughput scanner at an original resolution of 0.104 µm/pixel and subsequently downsampled to 0.5 µm/pixel for AI processing. Two SSL-based foundation models, Prov-GigaPath and UNI, were used to classify macrovesicular steatosis in frozen-section WSIs. Prov-GigaPath is a Vision Transformer-based model pretrained in a self-supervised manner on more than 170,000 WSIs from multiple tissue types.^22^ It employs dilated self-attention mechanisms to capture spatial relationships across entire slides, enabling robust recognition of histopathological features. UNI is a self-supervised pathology encoder trained on 100 million histopathology image patches and 100,000 WSIs, designed for multi-scale representation learning with strong transfer performance.^23^ From each WSI, model-derived embeddings were computed and used as inputs to downstream slide-level classifiers. A linear classifier was trained on these embeddings with class weights to address category imbalance, using permanent-section steatosis categories as labels.

### Model Training and Evaluation

Donors were randomly assigned 80:20 to a training cohort and a held-out test cohort. The held-out test cohort comprised 28 slides. Because the split was performed at the donor level, the number of slides per cohort does not necessarily follow an exact 80:20 ratio. All slides from the same donor were confined to a single cohort to prevent data leakage.

Within the training cohort, we performed four-fold cross-validation for model selection and hyperparameter tuning. Folds were created at the donor level with approximate balance of steatosis categories. After cross-validation, the linear classifier was refit on the full training cohort with the selected settings and then evaluated once on the held-out test cohort.

Performance was assessed for both the original five-category task and a binary task using the clinically relevant threshold of 30% (<30% vs ≥30%).

### Statistical Analysis

Interobserver agreement between two liver pathologists in quantifying macrovesicular steatosis was assessed using weighted Cohen’s kappa (κ) with quadratic weighting, accounting for the ordinal steatosis severity categories <5%, 5% to <10%, 10% to <20%, 20% to <30%, and ≥30%. Agreement between intraoperative frozen section diagnoses by on-call pathologists and final permanent section diagnoses was similarly evaluated using weighted Cohen’s kappa. Weighted kappa was calculated using the cohen_kappa_score function from the scikit-learn package (version 1.2) in Python 3.8. Prior to calculating weighted Cohen’s kappa, categorical labels were encoded numerically as 0 (<5%), 1 (5%–<10%), 2 (10%–<20%), 3 (20%–<30%), and 4 (≥30%). No missing values were present in the dataset; thus, no additional imputation was performed.

For comparative analysis, diagnostic accuracy of intraoperative frozen section diagnoses by on-call pathologists and AI models (Prov-GigaPath and UNI) was evaluated. Accuracy was assessed for both the five-category classification (steatosis severity categories) and a binary classification using a clinically relevant threshold of 30% (<30% vs. ≥30%). Performance was not stratified by individual pathologist training or years of experience due to the limited number of cases per reviewer; our aim was to assess overall diagnostic accuracy rather than subgroup differences. Statistical significance of accuracy differences between groups was determined using the chi-square test, performed in Python 3.8 (scipy.stats.chi2_contingency, SciPy version 1.10).

## Results

### Cohort Summary

The study cohort included 131 liver biopsy specimens obtained from 68 donors (31 men and 37 women). Macrovesicular steatosis distribution was as follows: <5% (n = 65; 49.6%), 5% to <10%, (n = 12; 9.2%), 10% to 20%, (n = 21; 16.0%), 20% to 30% (n = 8; 6.1%), and ≥30% (n = 25; 19.1%) (**Figure 2**). Interobserver agreement between the two liver pathologists was high (weighted Cohen’s kappa = 0.98), indicating strong consistency in steatosis assessment.

**Figure 2.**
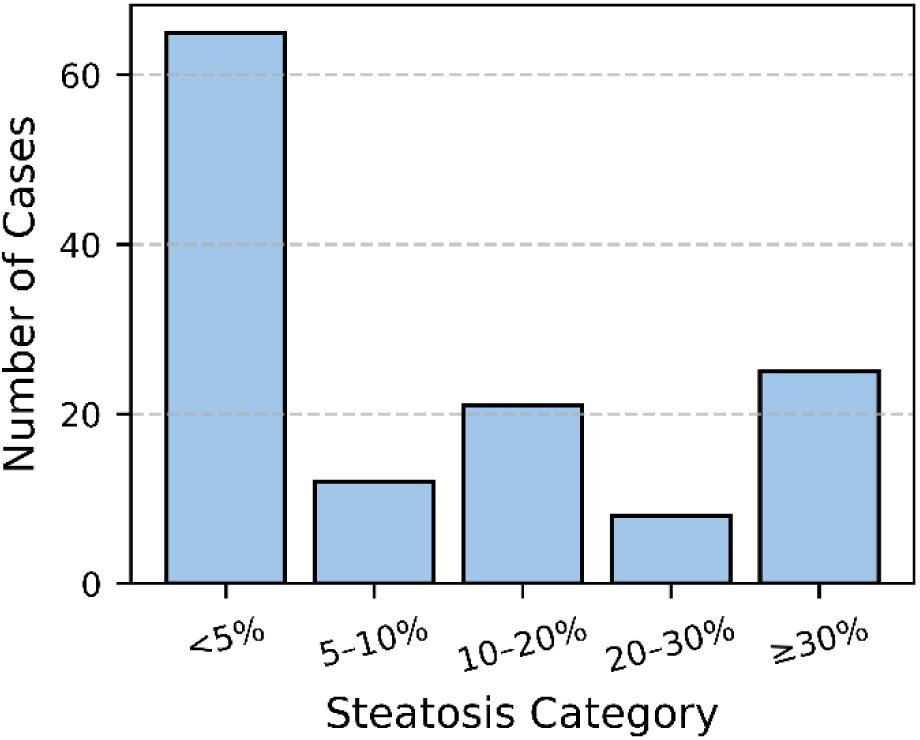
Distribution of macrovesicular steatosis in the study cohort. The histogram illustrates the distribution of 131 donor liver biopsies across five macrovesicular steatosis categories: <5%, 5% to <10%, 10% to <20%, 20% to <30%, and ≥30%.

### Intraoperative frozen and permanent sections

Among 131 donor liver biopsy specimens, intraoperative frozen section diagnoses showed an overall accuracy of 84.0% (110/131) when evaluated as a binary classification using a 30% steatosis threshold (<30% vs. ≥30%). Frozen section assessments correctly identified 95.2% (100/105) of specimens with <30% steatosis, but only 38.4% (10/26) of those with ≥30% steatosis.

When evaluating all five steatosis categories (<5%, 5% to <10%, 10% to 20%, 20% to 30%, ≥30%), frozen section diagnoses were concordant with permanent section diagnoses in 58.7% (77/131) of specimens, with a weighted Cohen’s kappa of 0.40, indicating fair agreement.

As shown in the confusion matrix (**Figure 3**), concordance was highest in the <5% steatosis category (87%, 57/65), whereas discrepancies occurred more frequently in categories ≥5%.

**Figure 3.**
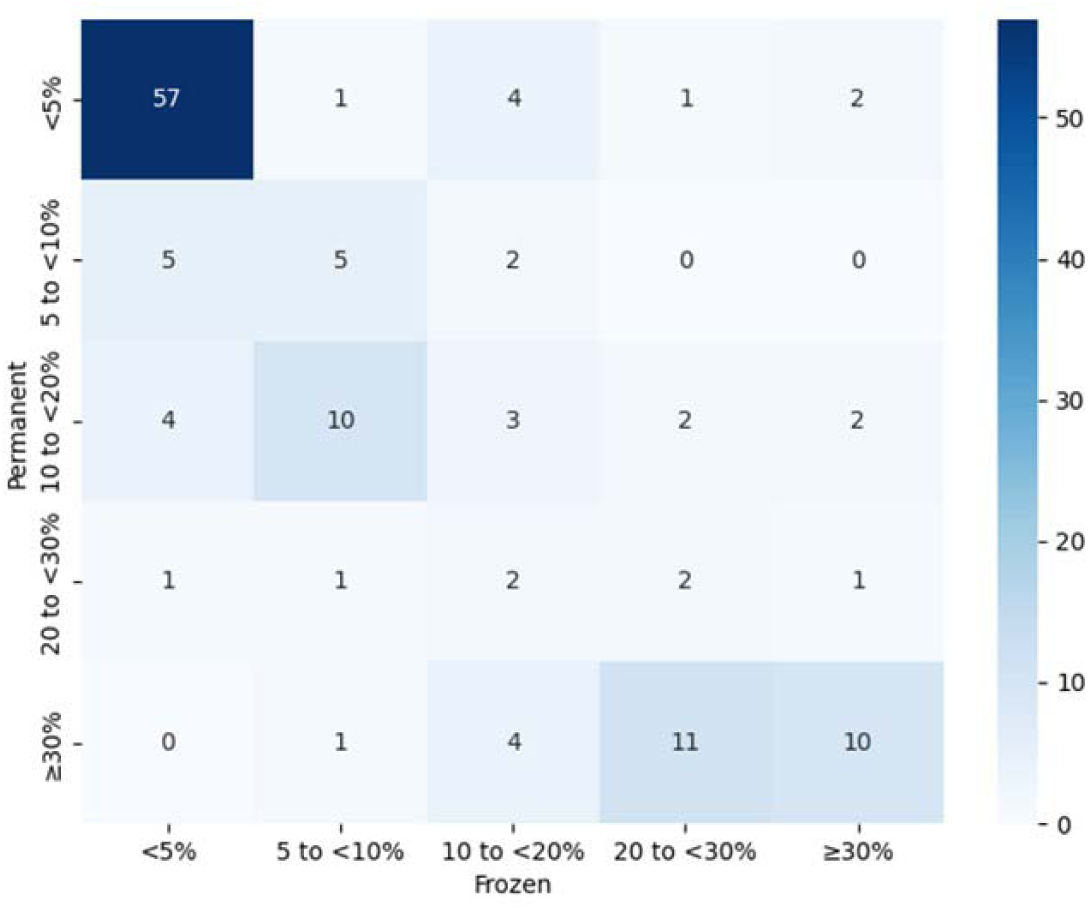
Confusion matrix of diagnostic discrepancies between frozen and permanent sections. The confusion matrix compares intraoperative frozen section diagnoses (x-axis) with ground truth permanent section assessments (y-axis). Color intensity indicates the number of cases in each category.

Notably, significant discrepancies included one specimen categorized as >30% steatosis by frozen section but subsequently found to have <5% steatosis on the permanent section (**Figure 4A, B**). Another specimen initially assessed as 5% to <10% steatosis intraoperatively was later categorized as >30% steatosis upon permanent section review (**Figure 4C, D**). These discrepancies likely arose from freezing artifacts, sampling variability, or inherent limitations of frozen section interpretation.

**Figure 4.**
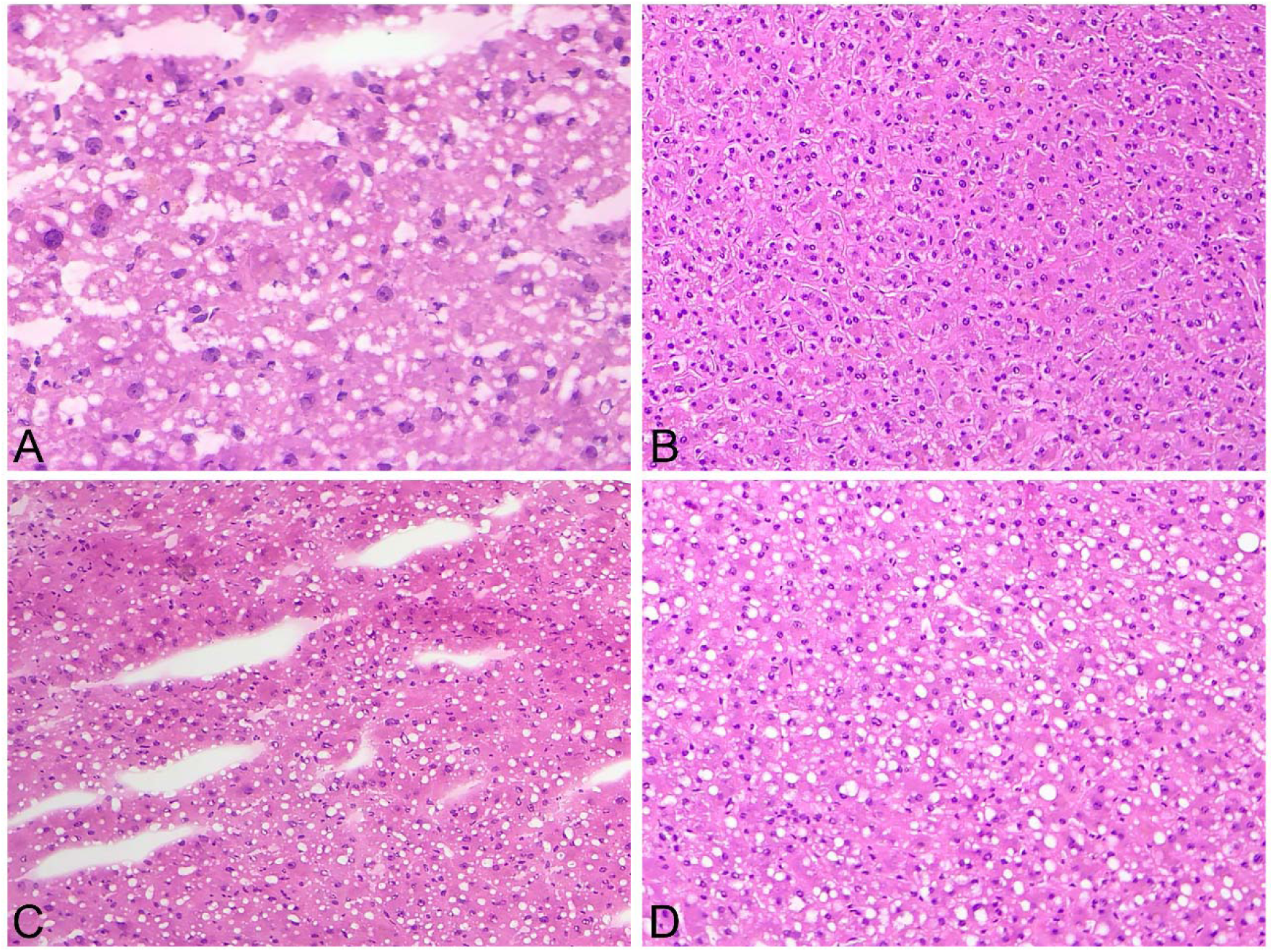
Representative cases of diagnostic discrepancies between frozen and permanent sections. (A, B) A donor liver biopsy is diagnosed as 50% steatosis on frozen section (A) but is reassessed as <5% on permanent section (B). (C, D) Another case is initially diagnosed as 5% to <10% steatosis on frozen section (C) but shows >30% steatosis on permanent section (D). All images are stained with hematoxylin-eosin, original magnification ×10.

### Prov-GigaPath model

In the training dataset, the Prov-GigaPath achieved an accuracy of 98.4% (129/131) in binary classification (<30% vs. ≥30%), correctly classifying 98.2% (109/111) with <30% steatosis and 100% (20/20) with ≥30% steatosis. For the five-category classification, overall accuracy was 88.4%. The model correctly identifying all 20 cases (100%) in the ≥30% category and 15 of 16 cases (93.8%) in the 10%–<20% category. Misclassifications were primarily observed in the <5% category, where 8 cases were incorrectly assigned to adjacent higher categories, indicating difficulty distinguishing minimal from mild steatosis (**Figure 5A**).

**Figure 5.**
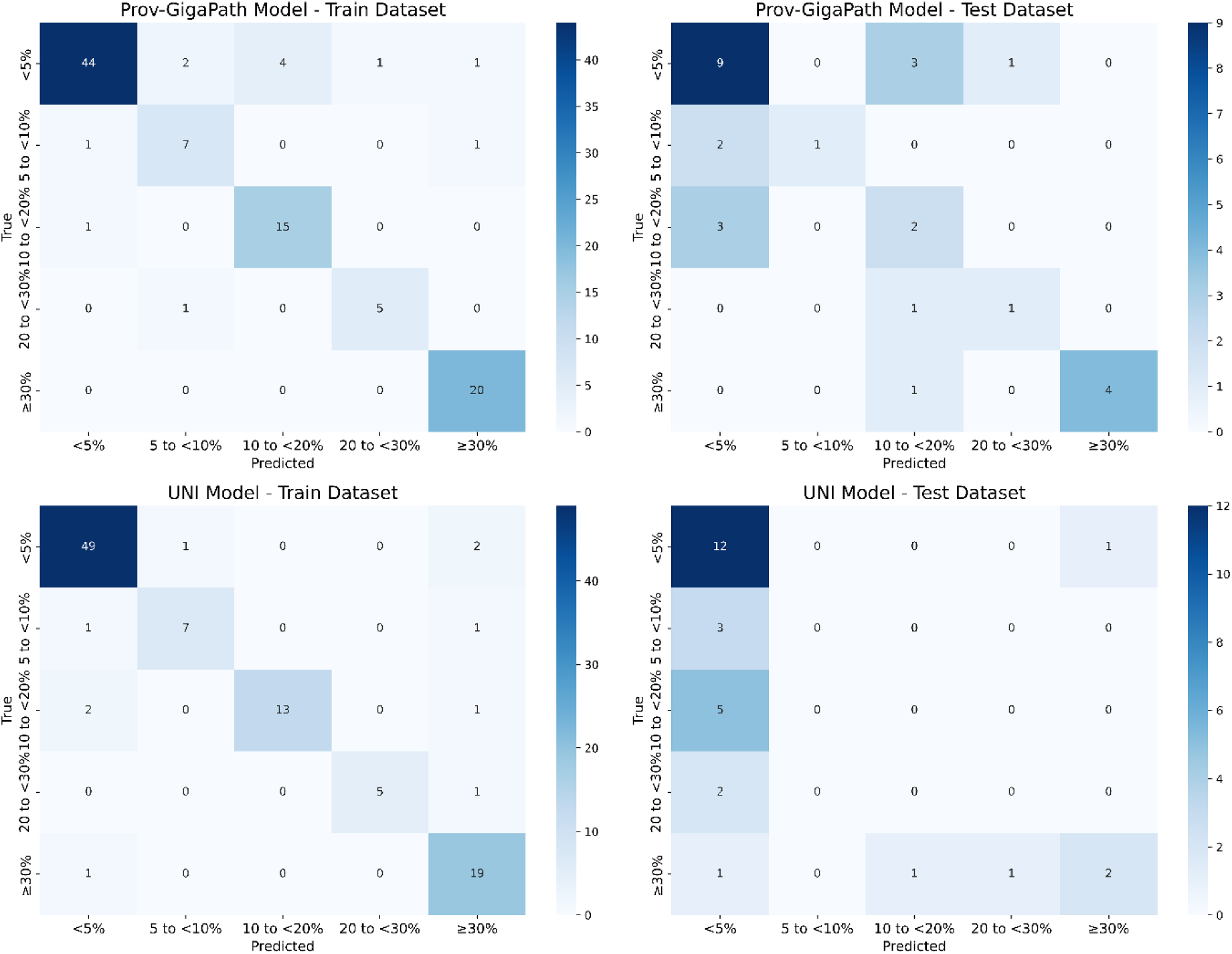
Confusion matrices for AI-based steatosis classification in frozen liver sections. The x-axis represents predicted categories, and the y-axis represents ground truth based on pathologist consensus. Color intensity indicates case count. (A) Prov-GigaPath model – training dataset. (B) Prov-GigaPath model – test dataset. (C) The UNI model – training dataset. (D) The UNI model – test dataset.

In the test dataset, the Prov-GigaPath achieved an accuracy of 96.4% (27/28 cases) in binary classification, correctly classifying all 23 cases with <30% steatosis and 4 of 5 cases (80.0%) with ≥30% steatosis. For the five-category classification, overall accuracy was lower at 57.1% (16/28). The model correctly classified 69.2% (9/13) of cases in the <5% category and 80.0% (4/5) in the ≥30% category. However, accuracy markedly decreased in the other three categories. Notably, the 10%–<20% category had no correctly classified cases, highlighting challenges in differentiating subtle differences in steatosis percentages under test conditions (**Figure 5B**).

### UNI Model

In the training dataset, the UNI achieved an accuracy of 96.9% (127/131) in binary classification (<30% vs. ≥30%), correctly classifying 96.4% (107/111) with <30% steatosis and 95.0% (19/20) with ≥30% steatosis. For the five-category classification, overall accuracy of 67.0%. The model performed well in classifying cases at the extremes of macrovesicular steatosis, correctly identifying 49 out of 52 cases (94.2%) in the lowest (<5%) and 19 out of 20 cases (95.0%) in the highest (>30%) steatosis categories. However, accuracy decreased in intermediate categories. The 10% to <20% steatosis group had 13 of 16 cases correctly classified, whereas the 5% to <10% and 20% to <30% categories exhibited more frequent misclassifications (**Figure 5C**).

In the test dataset, the UNI achieved an accuracy of 85.7% (24/28 cases) in binary classification, correctly classifying 95.7% (22/23) with <30% steatosis and 40.0% (2/5) with ≥30% steatosis. For the original five-category classification, overall accuracy was 50.0% (14/28). Accuracy was highest in the ≥30% category (80.0%, 4/5) and the <5% category (76.9%, 10/13), but substantially lower in the 5%–<10% and 10%–<20% categories, where no cases were correctly classified (**Figure 5D**).

Diagnostic accuracy of intraoperative frozen section assessment by on-call pathologists and AI models (Prov-GigaPath and UNI) is summarized in **Table 1**. In both binary and five-category classification tasks, accuracy differences among on-call pathologists and AI models were not statistically significant (binary classification: *P* = .22; five-category classification: *P* = .70).

**Table 1.** Comparison of diagnostic accuracy between on-call pathologists and ai models for steatosis classification

| **Classification method** | **On-call Pathologist** | **Prov-GigaPath** | **UNI** | **P value** |
| --- | --- | --- | --- | --- |
| Binary classification | 84.0% (110/131) | 96.4% (27/28) | 85.7% (24/28) | .22 |
| Five-category classification | 58.7% (77/131) | 57.1% (16/28) | 50.0% (14/28) | .70 |

Binary classification is based on a threshold of 30% steatosis (<30% vs. ≥30%). Five-category classification stratifies cases into five steatosis groups into five steatosis categories (<5%, 5% to <10%, 10% to <20%, 20% to <30%, and ≥30%). AI models are tested in 28 cases (approximately 20% of the total dataset) randomly selected as an independent test set. P-values are calculated using chi-square test.

## Discussion

Our study evaluated two self-supervised foundation models, Prov-GigaPath and UNI, for quantifying macrovesicular steatosis in donor liver biopsies. In the clinically critical binary classification (<30% vs. ≥30%), both models achieved high accuracy on the test dataset (Prov-GigaPath: 96.4%; UNI: 85.7%). Although their accuracy was numerically higher than those of intraoperative frozen-section assessments by on-call pathologists (84.0%), the differences were not statistically significant. Nevertheless, the high performance of these AI models highlights their potential for reliably identifying clinically relevant steatosis thresholds.

For the five-category steatosis classification, accuracy decreased for both AI models (Prov-GigaPath: 57.1%; UNI: 50.0%) and was numerically lower than that of intraoperative frozen section evaluations by on-call pathologists (58.7%), although this difference was not statistically significant. This lower accuracy was mainly due to the difficulty of differentiating subtle histological differences in underrepresented categories, such as 5% to <10%, 10% to 20%, and 20% to 30%. The dataset was imbalanced, with nearly half of samples (49.6%) in the <5% steatosis category and very few cases in other categories (e.g., only 6.1% in 20%–30%). Although SSL methods generally require fewer labeled data for fine-tuning than conventional supervised models, the extremely small sample sizes in these specific categories likely limited effective model training. Additionally, freezing artifacts common in intraoperative frozen sections may further complicate precise categorization at these intermediate levels. Given these findings, future implementation should prioritize optimized binary classification algorithms focusing on clinically significant thresholds (e.g., ≥30%) rather than multi-class categorization, which would enhance integration of AI into routine clinical decision-making.

Histopathological image analysis traditionally relied on CNN models, which have demonstrated robust performance in various pathology tasks, including steatosis quantification in liver biopsies.^26,27^ However, CNN-based models typically require substantial amounts of labeled data for effective training and fine-tuning, a significant limitation in pathology given the cost and complexity of obtaining expert-labeled histopathological datasets.^13,15^ In contrast, SSL-based foundation models, such as Prov-GigaPath and UNI, address this limitation by leveraging large-scale pretraining on extensive, unlabeled histopathological image repositories, enabling rapid adaptation to specific tasks with substantially fewer labeled samples.^22,23^ Recent comparative studies, such as the evaluation of the UNI foundation model, reported that UNI outperformed ResNet-50 and other pretrained encoders on average, with the greatest gains in rare cancer classification and complex diagnostic tasks.^28^ Our results support these findings, highlighting SSL-based foundation models’ capacity for effective feature extraction and generalization, which is particularly in settings where labeled data are limited.

These models inherently capture broad morphological features and context across WSIs, an advantage over traditional CNN methods that typically analyze small local patches and may overlook spatially relevant histological features. The transformer architecture, employed by Prov-GigaPath, further enhances this global feature integration by utilizing self-attention mechanisms to better preserve spatial context across gigapixel images.^29,30^ Collectively, these strengths position SSL-based foundation models as a highly attractive approach for clinical deployment, enabling precise histopathological assessments from relatively limited annotated data.

The integration of WSI into intraoperative frozen section evaluation presents several practical challenges. A key limitation is the additional scanning time required, as even a few extra minutes per slide could disrupt workflow efficiency in the time-sensitive setting of liver transplantation. While scanning technology continues to improve, current turnaround times remain suboptimal for real-time decision-making. In our study, all 131 frozen-section slides were digitized in a single batch, and because we did not record individual scan or inference times, we cannot report exact single-slide turnaround, which limits the quantitative assessment.

Additionally, pathology residents or assistants responsible for frozen section preparation would need training to operate scanners, introducing workflow adjustments and potential variability. Further studies should assess the feasibility, time efficiency, and cost-effectiveness of incorporating WSI and AI-based analysis into routine intraoperative workflows.

Another limitation is the modest size of our independent test set (nD=D28), which limits statistical power to detect differences between AI models and pathologists. In addition, the uneven distribution of steatosis categories, predominantly <5%, limited AI model performance in intermediate categories. Our analysis was also focused solely on classical macrovesicular steatosis, and we did not evaluate small-droplet macrovesicular steatosis or microvesicular steatosis due to the lack of standardized definitions.^3^ Consequently, the AI models may misclassify these patterns, representing a limitation of the current study. Future research should include balanced datasets that represent these lipid patterns, employ advanced data augmentation techniques, and integrate other histologic parameters such as inflammation and fibrosis. Addressing these elements would enhance the generalizability of AI models and provide a more comprehensive and clinically relevant assessment for transplant.

Finally, this study did not include transplant recipient outcome data, as intraoperative frozen section evaluations were performed in collaboration with the Gift of Life donor program, which does not provide post-transplant follow-up. Our study compared AI models and pathologists in assessing macrovesicular steatosis but adding transplant outcomes would increase clinical relevance. Ideally, AI models should be trained to predict graft survival and patient prognosis, which would help guide transplant decisions.

In conclusion, our study supports the clinical utility of SSL-based foundation models for reliable quantification of macrovesicular steatosis in donor liver biopsies. While limitations remain, particularly regarding intermediate steatosis categories and clinical integration challenges, the demonstrated advantages in accuracy, label efficiency, and generalization highlight their promising potential. Further refinement of training methods and inclusion of clinical outcome data could advance AI-based steatosis evaluation toward standardized, objective intraoperative assessments that improve transplant decision-making and patient outcomes.

## Ethical Approval

This study was approved by the Institutional Review Board at the Hospital of the University of Pennsylvania, meeting exemption criteria (category 4) with a HIPAA waiver. This study was conducted in accordance with the Declaration of Helsinki 1975.

## Informed Consent

In accordance with institutional policy, the hospital obtains general consent at the time of care that permits secondary research use of de-identified clinical data and archived pathology material; therefore, the Institutional Review Board waived the requirement for study-specific informed consent.

## Data Availability Statement

All data produced in the present study are available upon reasonable request to the authors.

